# Heat stress-induced heat shock protein 70 (HSP70) expressions among vulnerable populations in urban and rural areas Klang Valley, Malaysia

**DOI:** 10.1101/2023.12.20.23300317

**Authors:** Siti Nurfahirah Muhamad, Abdah Md Akim, Fang Lee Lim, Nur Shabrina Azreen Mohd Shabri, Vivien How

## Abstract

As climate change raises global temperatures, there remains a notable gap in understanding the body’s mechanism of the heat stress defense exhibited by Heat Shock Protein (HSP) within the populations. This study aims to evaluate the expression of heat shock protein 70 (HSP70) in vulnerable populations living in urban and rural areas in response to heat exposure. A comparative cross-sectional study involved 54 urban and 54 rural participants from Klang Valley, Malaysia. This study comprises four methods: Part I involves conducting face-to-face interviews questionnaire; Part II involves monitoring indoor heat exposure and classifying thermal stress using the Universal Thermal Climate Index (UTCI); Part III involves using the reverse-transcription quantitative polymerase chain reaction (RT-qPCR) and Part IV involves using the HSP70 High Sensitivity Enzyme-linked Immunosorbent Assay (ELISA). The findings revealed that urban areas have a higher heat level, classified as strong UTCI thermal stress (32.1°C), whereas rural areas have moderate UTCI thermal stress (31.0°C). In response to heat stress, the urban vulnerable populations exhibited higher HSP70 gene expression (0.167 ± 0.86) compared to the rural (0.154 ± 0.28). A significant difference (*p*<0.001) in HSP70 protein expression was observed in the plasma of urban compared to rural vulnerable populations. There was a strong association between UTCI heat exposure level and the expression of the HSP70 gene and protein in both vulnerable population groups (*p*<0.001). Although susceptible to vulnerabilities, the populations demonstrated HSP70 expressions in response to varying levels of heat exposure as a coping mechanism at the cellular level.

## Introduction

Climate change, driven by the increase in average global temperatures due to the release of greenhouse gases, poses significant adverse effects [1]. Methane, chlorofluorocarbon, and nitrous oxide are examples of greenhouse gases that contribute to the greenhouse effect. These gases trap solar radiation, leading to elevated global temperatures and changes in weather patterns. As a result, the frequency and intensity of heat waves are anticipated to increase. Heatwaves are extreme heat events that profoundly impact the environment, infrastructure, human health, and social well-being [2].

Urban areas characterized by high population density and unanticipated climate variations are prone to the Urban Heat Island (UHI) phenomenon and heightened heat exposure. Malaysia is also affected by this phenomenon, as seen in the increasing trend of daily temperatures over the past decade [3, 4]. Excessive heat exposure can lead to heat stress, heat-related illnesses such as heat stroke, an elevated risk of respiratory, cardiovascular, and renal issues, and unfavorable perinatal outcomes [5-7]. However, the risk of these adverse effects can be mitigated through heat acclimatization, a process in which repeated exposure allows the body to adjust and sustain optimal performance in varying circumstances [7, 8].

The physiological response that ensues to maintain an optimal core body temperature, such as heat dissipation and the activation of HSPs, specifically HSP70, play a vital role in thermotolerance. The HSP70 gene and protein are constantly expressed at essential levels in normal physiological conditions as a part of protein homeostasis [9, 10]. When the body is exposed to elevated temperature, heat shock factors (HSFs) will be activated and attach to the HSP70 gene to form heat shock elements (HSEs), ultimately leading to the transcription of the HSP70 gene [9]. It would later translate into protein to facilitate the cellular response, safeguarding the cell from potentially harmful high temperatures that could cause damage to the cell [11].

Among the family of HSPs, HSP70 is the most thermally sensitive and functions as a cellular thermometer. It reacts to heat stress and other stimuli to support cellular processes, promote cell survival, and prevent apoptosis [9, 12-15]. Dysregulation of thermal control can cause cellular damage, hypercoagulation with suppressed fibrinolysis, and hyperinflammation, ultimately resulting in the lethal consequences of heat stroke [16-17]. Therefore, understanding the dynamics of HSPs, especially HSP70, is essential for comprehending the body’s defense mechanisms against heat-related adverse effects.

Previous studies have provided increasing evidence that vulnerable populations are the most affected by heat-related mortality and morbidity impacts [5, 18-20]. Due to the disproportionate effects of climate change on different vulnerabilities, such as the low-income, elderly, minorities, and those with chronic health conditions [21, 22], previous studies have primarily focused on urban areas, overlooking the vulnerabilities and adaptations of rural communities. Therefore, this study aims to investigate the expression level of HSP70 to heat exposure among vulnerable populations in urban and rural environments. This research seeks to provide valuable insights into the adaptive capabilities of different populations in dealing with heat-related adverse effects. The findings will serve as a baseline for improving strategies to mitigate the severity of health impacts associated with climate change.

## Materials and methods

### Participants

A comparative cross-sectional study was conducted among 108 Malaysians aged 13 and above. The data was collected from 28 July to 23 September 2022 during the Southwest Monsoon in Malaysia. A cluster random sampling method was used to select four urban and four rural areas in the Klang Valley regions. Subsequently, a stratified random sampling method was utilized to ensure an equal number of respondents from both urban and rural areas. The study included individuals from vulnerable groups, including senior citizens (aged 60 years or older), low-income individuals (those in B40 groups who represent the bottom 40% of income in Malaysia), and individuals with health morbidities, which live in their current residential areas more than one year. On the other hand, pregnant women and individuals under 13 years were excluded from this study. Written consent was obtained from participants and parents or guardians for minors before proceeding with the data and samples collection. During the data collection, several methods were conducted, including a face-to-face questionnaire, indoor heat exposure monitoring and thermal stress classification using the UTCI, and blood sample collection for RT-qPCR and ELISA analysis. The data collection took place in a designated room located within the participants’ residential areas to authentically represent the conditions of their living environment.

### Face-to-face questionnaire

A validated questionnaire was used to obtain the sociodemographic background and health status of the study population. The data was obtained through face-to-face interviews, and only participants who met the inclusive criteria were selected for the study.

### Indoor heat exposure monitoring

The designated area (sampling area) was arranged to operate without the use of any ventilation systems, such as fans and air conditioning (AC). Only windows were opened for natural ventilation. Indoor heat exposure was monitored using a Wet-bulb Globe Temperature (WBGT) monitor (Model: QUESTemp). The WBGT monitor was placed at a height of 1.1 meters from the floor level, where the heat stress is at its maximum, based on the recommendation in ISO 7243 [23]. The reading was recorded during the mid-day hours between 12 noon and 3 p.m. for one hour to measure four parameters simultaneously, which are ambient temperature, radiant temperature, relative humidity, and air velocity at the same time. The data obtained from WBGT was calculated using the formula of the Universal Thermal Climate Index (UTCI) [24] as follows:

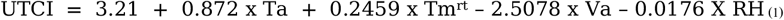

where T_a_ is air temperature, T_mrt_ is mean radiant temperature, V_a_ is wind speed, and RH is relative humidity. The calculated value was then used to determine the respondents’ heat exposure level and thermal stress category based on the UTCI equivalent temperature scale [25].

### Blood sample collection for RT-qPCR and ELISA analysis

After the participants were exposed to the designated sampling area indoor temperature for 30 minutes, a total of 5ml blood sample was collected from each participant via venipuncture blood procedure into two Ethylenediaminetetraacetic acid (EDTA) tubes. Tube A (2ml) was used for RT-qPCR, and Tube B (3ml) was used for ELISA analysis.

### Reverse-transcription Quantitative Polymerase Chain Reaction (RT-qPCR)

RT-qPCR was conducted to determine the level of HSP70 gene expression. This method involves RNA isolation, cDNA conversion, and quantitative PCR (qPCR). A total of 1.5ml blood from Tube A was used to extract the total RNA using the QIAamp RNA Blood Mini Kit (Model: Qiagen) within 24 hours after the blood samples were collected, following the manufacturer’s procedures [26]. The purity of the extracted RNA was assessed by spectrophotometry, and the RNA integrity was examined using gel electrophoresis before proceeding with cDNA conversion procedures. The cDNA synthesis used the QuantiTech Reverse Transcription Kit (Model: Qiagen).

Quantitative PCR (qPCR) provides quantification of HSP70 gene expression. qPCR was conducted using Quantinova SYBR Green PCR kit (Model: Qiagen) analyzed in the PCR cycler (Model: Mastercycler® ep Realplex 4, Eppendorf, Germany). The three steps of thermal cycling begin with PCR initial activation step (2 mins, 95°C), followed by 2-step cycling of denaturation (5secs, 95°C), and combined annealing or extension (10 secs, 60°C) for a total of 40 cycles. A standard curve was performed to obtain the PCR efficiency for the primers of the HSP70 gene (NM_005527.3) and 18SrRNA (housekeeping gene) [27].

The Ct-values of experimental samples obtained from the qPCR analysis were calculated using relative quantification to get the level of HSP70 gene expression [28]. The primers’ sequences are 5’TTCGTGGCTGGAGGTCAATC-3’, 5’-TAATGATTTGAAGATGAGGGG-3’ (HSP70 gene) and 5’-GTGGTGTTGAGGAAAGCAGACA-3’, 5’-TGATCACACGTTCCACCTCATC-3’(18SrRNA) [27]. The average Ct-values of HSP70 and 18srRNA genes for each sample obtained from the qPCR procedure were calculated in the linear equation generated from the standard curve. The computed value of the HSP70 gene was then normalized to the 18srRNA gene value to obtain the HSP70 gene expression (expressed in ratio value). The normalization was done to ensure that the quantified HSP70 gene represents heat-induced expression.

### HSP70 high-sensitivity Enzyme-linked Immunosorbent Assay (ELISA)

HSP70 protein expression was measured using a sandwich method HSP70 high-sensitivity enzyme-linked immunosorbent assay (ELISA) Kit (Model: Abcam, ab133061). The blood sample in Tube B was used to analyze HSP70 protein expression levels. The plasma was separated by centrifugation at 1000 x g for 15 minutes at 4°C and prepared with a dilution of 1:4 in the assay buffer. HSP70 concentration levels were expressed in ng/ml. The assay’s sensitivity or detection limit is 0.09 ng/mL (90 pg/mL). Quantitative determination of HSP70 in the plasma was performed based on a standard curve [29]. The assayed ELISA plate was immediately analyzed using a microplate reader (Model: Biotek 800 TS Absorbance Reader). The optical density (OD) absorbance measurement was recorded at 450nm. The results were then calculated using a standard curve generated using 4PL equations as recommended by the manufacturer’s protocol [30].

### Statistical analysis

The data obtained from the respondents were analyzed using Statistical Package for the Social Sciences (SPSS) version 25. The comparison between UTCI heat exposure level, HSP70 gene, and HSP70 protein expressions was analyzed using the Independent t-test or Mann-Whitney U Test. A correlation test was conducted to determine the association between the variables. *P*-values less than 0.05 (*p*<0.05) were considered significant.

## Results

### Characteristics of the participants

This study involved 108 participants, with 54 respondents from urban and 54 from rural areas. Urban respondents comprised 27.8% senior citizens (60 years old and above), 90.7% B40 groups (low-income people), and 46.3% people with health morbidity. On average, the participants live in their residential areas about 16.6 ± 6.34 years.

In rural, there are 29.6% senior citizens, 88.9% B40 groups, and 53.7% people with health morbidity. On average, the participants live in their residential areas about 30.4 ± 6.52 years. Table 1 shows the sociodemographic characteristics and health status of the study population.

**Table 1.** Sociodemographic Characteristics and Health Status of the Study Population (N=108)

| Continuous variables | Urban (n=54) | Rural (n=54) |
| --- | --- | --- |

|  | Mean (SD) |  |
| --- | --- | --- |
| Age (years) | 52.8 (11.44) | 49.4 (16.1) |
| Body mass index, BMI (kg/m<sup>2</sup>) | 28.9 (6.99) | 27.2 (6.42) |
| Duration of residency (years) | 16.6 (6.34) | 30.4 (6.52) |
| **Categorical variables** | **Frequency, *f* (%)** |  |
| Age groups |  |  |
| <60 years old | 39 (72.2) | 38 (70.4) |
| ≥60 years old | 15 (27.8) | 16 (29.6) |
| Gender |  |  |
| Male | 8 (14.8) | 18 (33.3) |
| Female | 46 (85.2) | 36 (66.7) |
| Educational level |  |  |
| No formal education | 7 (13.0) | 1 (1.6) |
| Primary school | 6 (11.1) | 16 (29.6) |
| Secondary school | 37 (68.5) | 27 (50.0) |
| Diploma/STPM/Matriculation | 4 (7.4) | 5 (9.3) |
| Bachelor | 0 (0.0) | 5 (9.3) |
| Household income (RM) |  |  |
| B40 (Below RM4851) | 49 (90.7) | 48 (88.9) |
| M40 (RM4851-RM10,970) | 5 (9.3) | 6 (11.1) |
| Health morbidity |  |  |
| Yes | 25 (46.3) | 29 (53.7) |
| No | 29 (53.7) | 25 (46.3) |

Abbreviations: SD, standard deviation; RM, Ringgit Malaysia

### UTCI heat exposure level

The average UTCI heat exposure level for urban areas is 32.1°C, classified as strong thermal stress. For rural areas, the average UTCI heat exposure level was 31.0°C, classified as moderate thermal stress. There was a significant mean difference in UTCI heat exposure level between urban and rural areas (*p*<0.001). The results indicated that urban people were exposed to higher heat stress than rural people. Fig 1 shows the UTCI heat exposure level between urban and rural areas.

**Fig 1.**
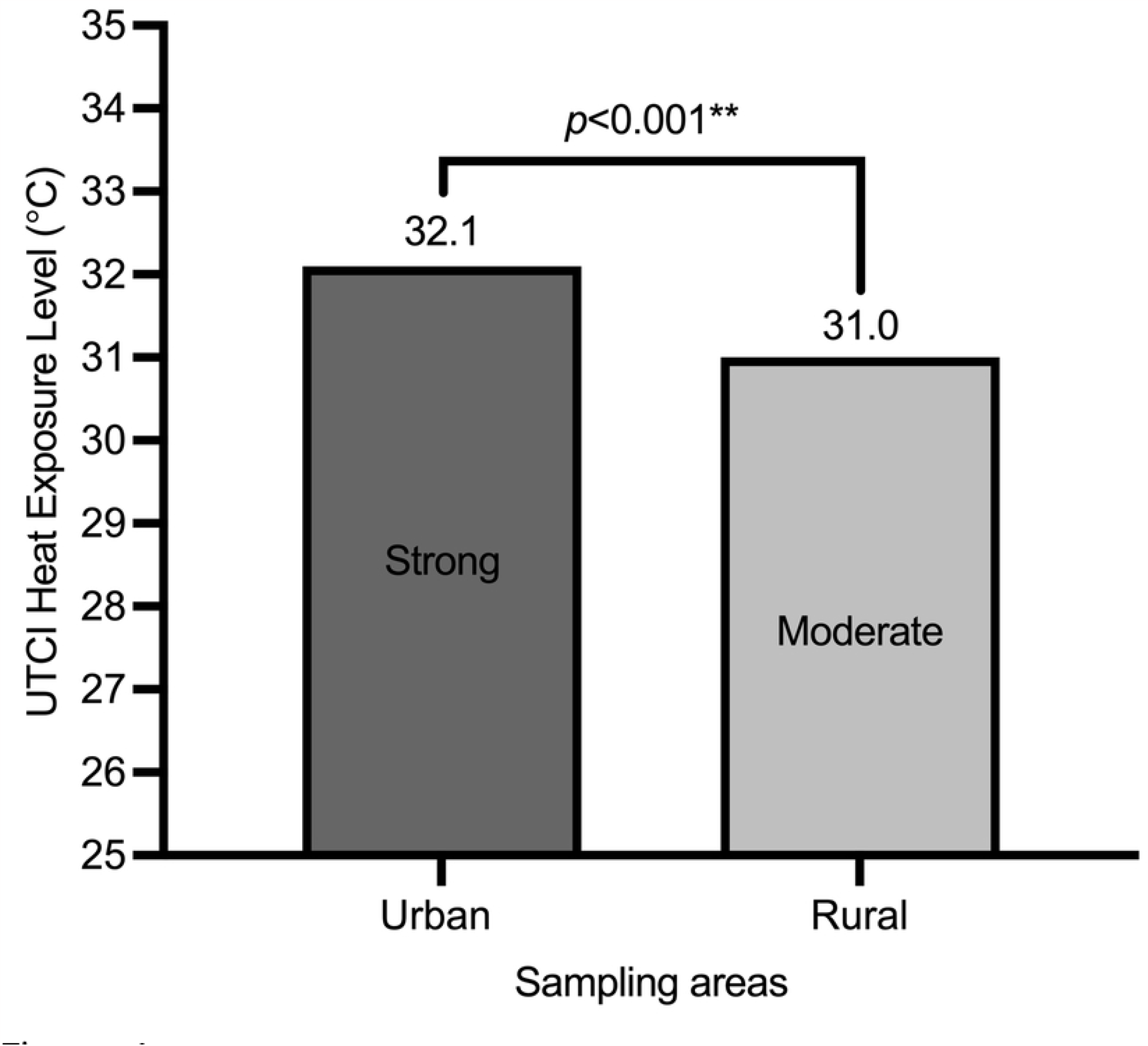
UTCI Heat Exposure Level between Urban and Rural Areas (N=108)

### HSP70 gene expression

On average, urban recorded higher HSP70 gene expression (0.167 ± 0.86) than rural vulnerable populations (0.154 ± 0.28). However, there was no significant difference in HSP70 gene expression between urban and rural vulnerable people (*p*>0.05). Fig 2 shows the distribution of HSP70 gene expression between both groups.

**Fig 2.**
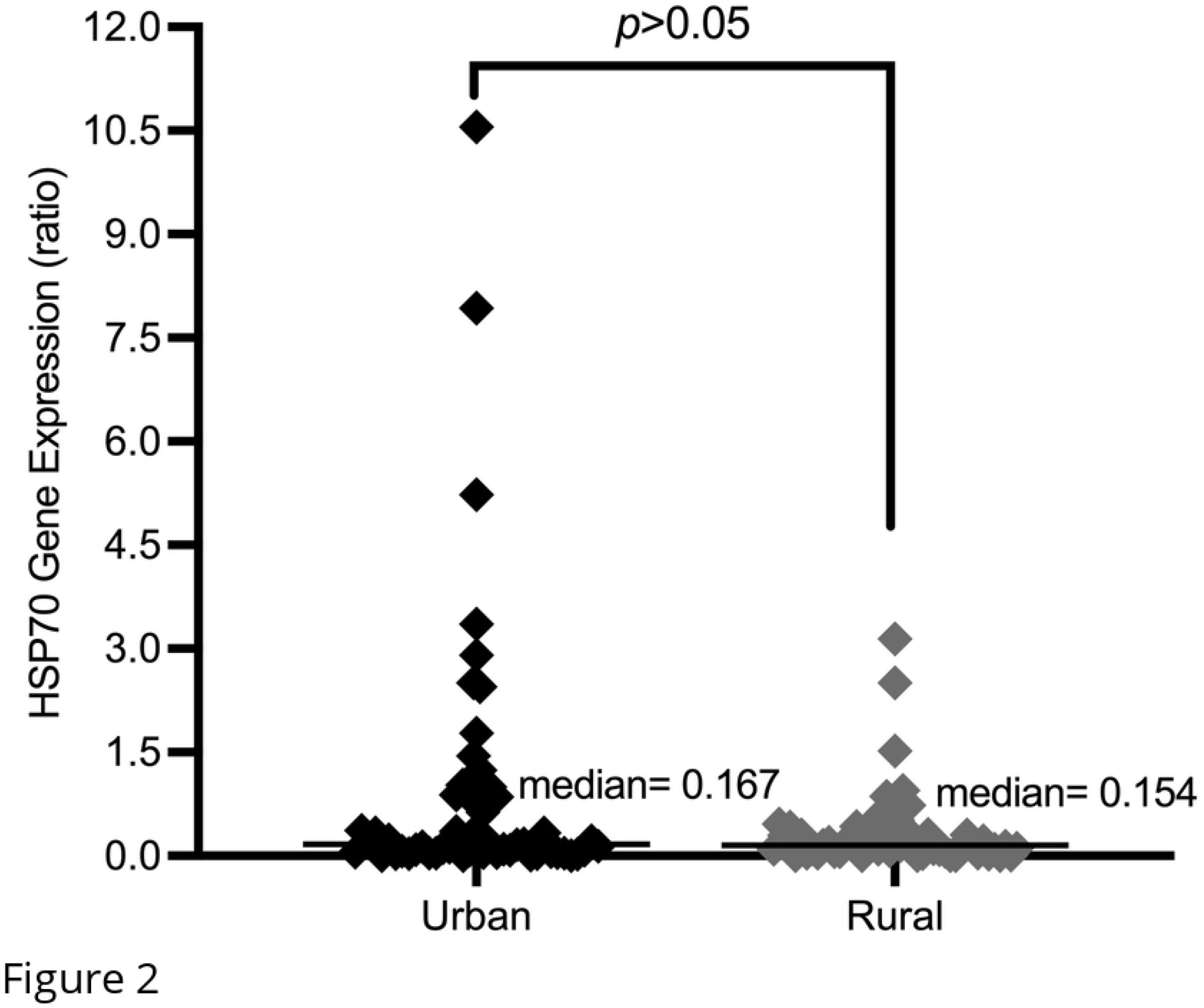
Distribution of HSP70 Gene Expression between Urban and Rural Vulnerable Populations (N=108)

### HSP70 protein expression

Urban demonstrated a higher HSP70 protein expression (2.524 ng/ml) than rural vulnerable populations (2.095 ng/ml). There was a significant mean difference in HSP70 protein expression between urban and rural vulnerable groups (*p<*0.001). Fig 3 shows the distribution of HSP70 protein expression between both groups.

**Fig 3.**
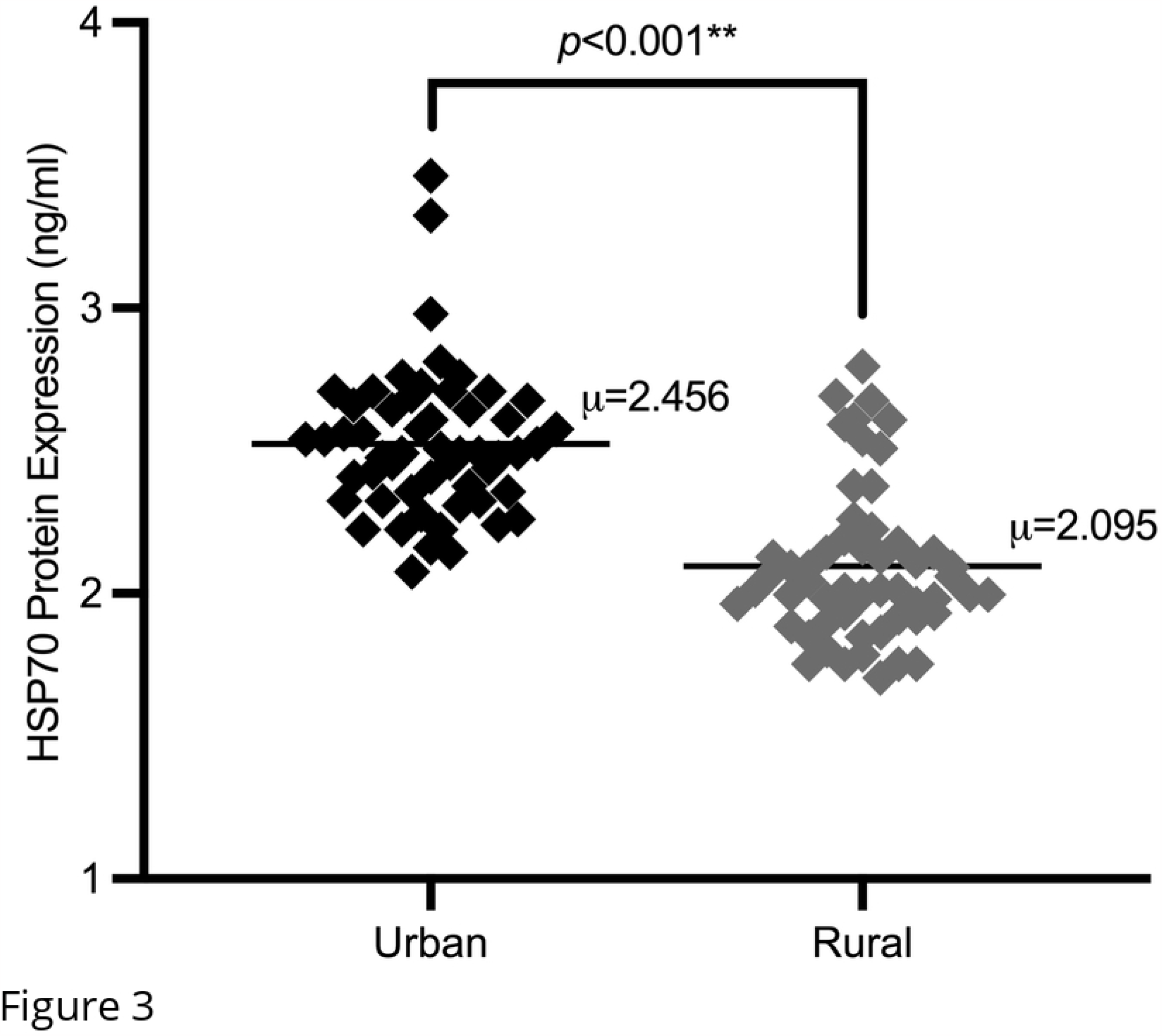
Distribution of HSP70 Protein Expression between Urban and Rural Vulnerable Populations (N=108)

### Association between UTCI heat exposure level with HSP70 expressions

A correlation test was done to determine the association between UTCI heat exposure level and HSP70 expressions. For urban areas, there was a significant positive correlation between UTCI heat exposure level with HSP70 gene expression (r = 0.592, *p*<0.001) and HSP70 protein expression (r = 0.392, *p*<0.001). Table 2 shows the association between UTCI heat exposure level and HSP70 expressions for urban vulnerable populations.

**Table 2.** Association between UTCI Heat Exposure Level and HSP70 Expressions for Urban Vulnerable Populations (n=54)

Statistical test: <sup>a</sup> Pearson correlation; <sup>b</sup> Spearman rho
| Variables | Heat exposure level<sup>a</sup> | HSP70 gene expression<sup>b</sup> | HSP70 protein expression<sup>a</sup> |
| --- | --- | --- | --- |
| Heat exposure level<sup>a</sup> | 1 |  |  |
| HSP70 gene expression<sup>b</sup> | 0.592** | 1 |  |
| HSP70 protein expression<sup>a</sup> | 0.392** | 0.395** | 1 |

\* Correlation is significant at the 0.05 level (2-tailed)
\*\* Correlation is significant at the 0.01 level (2-tailed)

For rural areas, there was a significant positive correlation between UTCI heat exposure level with HSP70 gene expression (r = 0.578, p<0.001) and HSP70 protein expression (r = 0.360, *p*<0.001). Table 3 shows the association between UTCI heat exposure level and HSP70 expressions for rural vulnerable populations.

**Table 3.** Association between UTCI Heat Exposure Level and HSP70 Expressions for Rural Vulnerable Populations (n=54)

| Variables | Heat exposure level<sup>a</sup> | HSP70 gene expression<sup>b</sup> | HSP70 protein expression<sup>a</sup> |
| --- | --- | --- | --- |
| Heat exposure level<sup>a</sup> | 1 |  |  |
| HSP70 gene expression<sup>b</sup> | 0.578** | 1 |  |
| HSP70 protein expression<sup>a</sup> | 0.360** | 0.128 | 1 |

Statistical test: <sup>a</sup> Pearson correlation; <sup>b</sup> Spearman rho
\* Correlation is significant at the 0.05 level (2-tailed)
\*\* Correlation is significant at the 0.01 level (2-tailed)

## Discussion

Expression of the HSP70 gene and protein synthesis are tightly regulated and play a vital role in cellular homeostasis during heat stress [31]. This study found that urban respondents had higher levels of HSP70 gene and HSP70 protein expression compared to rural respondents. This finding is consistent with the recorded heat exposure level in both areas, where urban areas recorded higher heat exposure than rural areas. A significant mean difference in HSP70 protein expression was observed between individuals residing in urban and rural areas (*p*<0.001). This finding aligns with previous studies that have documented a significant difference in the HSP70 protein expression at different levels of thermal exposure [32, 33].

However, there was no significant difference in HSP70 gene expression between urban and rural respondents (*p*>0.05). The finding can be explained by the observation that individuals living in urban areas exhibited only a marginally elevated HSP70 gene expression compared to individuals in rural areas. Since the result did not reflect a similar trend of the HSP70 protein, it can be inferred that the cellular response primarily takes place at the protein level. Even though the expression of the HSP70 gene plays a role in stress-coping mechanisms, the study suggests that its effects in this context are relatively modest. This finding can be elucidated by the fact that individuals who have adapted to high temperatures have a substantially elevated baseline level of HSP70 protein [34]. Several analyses comparing HSP70 gene and protein measurements following heat acclimation found an inconsistency, suggesting that an increase in HSP70 protein levels may not correspond to an increase in HSP70 gene levels. In fact, another study revealed that once the response requirements of HSP70 are met, the previous built-up of HSP70 protein levels may interact with Heat Shock Factor 1 (HSF-1) to further inhibit the transcription of the HSP70 genes [11].

The primary mechanism for negative feedback control involves the binding and decoupling of HSP70 with HSF (HSP: HSP Complex), where the phosphorylation would later lead to amplifying the transcriptional output [35]. Protein degradation (such as unfolded or denatured proteins) increases as the temperature rises. This condition induces the separation of chaperone HSP70 from HSF-1, allowing HSF-1 to stimulate the production of more HSP70 and increase the transcription of the HSP70 gene [36]. Nevertheless, sufficient HSP70 protein is necessary to reinstate proteostasis and prevent cellular stress. The adequate HSP70 protein copes with the cellular stress, facilitating the binding between HSP70 and HSF-1, leading to a decrease in the phosphorylation and transcription of the HSP70 gene. Although urban respondents experience higher levels of heat exposure and have significant differences in HSP70 protein, their level of HSP70 gene expression indicates that they are tolerant to the heat they are exposed to. In other words, urban respondents have sufficient HSP70 protein to maintain the proteostatis, which leads to the repression of HSF-1 and prevents the activation and induction of new HSP70 gene expression.

In this study, urban and rural respondents showed a significant association between UTCI heat exposure level and HSP70 expressions. These findings align with previous research that demonstrated thermal stress can induce HSP70 protein expression [11, 37] and HSP70 gene expression [33, 34]. The previous studies maintained a constant ambient temperature at 40°C (which exceeds the normal human physiological body temperature range) coupled with a relative humidity of 20-45% and found an upregulation in the expression of the HSP70 gene [33, 38] and HSP70 protein [34, 39] in leukocytes. In contrast, findings from this study demonstrate a disparity in the expression of the HSP70 gene and protein in leukocytes between urban and rural respondents, even when exposed to a temperature of 29-31°C (ambient temperature) with relative humidity of 69-75%.

While the respondents in this study experienced a lower ambient temperature than the designated exposure level in the previous studies [33, 34, 38, 39], the elevated relative humidity in Malaysia, known for its hot and humid climate, could greatly affect the overall thermal stress experienced. This finding is supported by a previous study, which stated that high temperatures (30-40°C) and high humidity (>60%) can increase the risk of heat-related illness and significantly trigger the body’s thermoregulatory response [40]. Furthermore, the heat shock response at the cellular level is a dynamic and adaptive process that is not dependent on reaching the specific temperature threshold [41]. In fact, it could also be altered by other factors such as exposure duration, intensity, and acclimation to heat [11, 33, 42]. This study revealed that vulnerable populations living in hot and humid tropical areas are capable of inducing HSP70 expressions as a means of adapting and coping with heat stress.

## Conclusion

Overall, this study found that the average UTCI heat exposure level in urban areas was higher than in rural areas. Although experiencing increased heat stress, vulnerable people in urban demonstrated better heat tolerance compared to rural people based on their HSP70 gene and protein patterns as coping and adapting to heat stress. This study also suggests that individuals in both urban and rural areas susceptible to heat stress in hot and humid regions have exhibited cellular responses. Therefore, these recent findings are favorable for addressing the necessary heat mitigation and adaptation responses following the highlighted heat exposure levels and cellular responses in urban and rural areas. However, different climates may exhibit distinct heat stress responses due to the potential influence of heat acclimatization on the outcome. Therefore, further investigation of the heat stress response in vulnerable populations is recommended in comparing tropical and temperate climate regions.

## Data Availability

All relevant data are within the manuscript and its Supporting Information files.

## Acknowledgment

The authors thank Mrs. Amrina Mohd Amin and Mrs. Safarina Mohamad Ismuddin for their assistance during the field sampling and laboratory work.

